# Assessing the carbon footprint of clinical trials: a systematic review

**DOI:** 10.1101/2024.11.12.24317142

**Authors:** Charline Jean, Richard Layese, Florence Canouï-Poitrine, David Grimaldi, Etienne Audureau, Michelle Leemans, Charlotte Lafont

## Abstract

**Background:** The healthcare sector substantially contributes to global greenhouse gas emissions. While being pivotal for improving care, clinical trials involve various activities beyond routine care that contribute to their carbon footprint. We aimed to synthesize current evidence on the carbon footprint of clinical trials and the methodologies used to estimate these emissions.

**Methods:** In this systematic review, we searched PubMed, Embase, and Cochrane databases for studies published in English until April 16, 2024. Studies focusing on the carbon footprint of clinical trials were included. Abstracts without full-text availability were excluded. Four reviewers independently extracted data, focusing on trial characteristics, carbon emission quantification methodologies, and reported emissions per trial and patient. The risk of bias was assessed using a transparency checklist for carbon footprint calculations.

**Findings:** The review included 12 studies (6 analytical studies and 6 expert opinions). Total emissions per trial varied widely, ranging from 18 to 2,498 tons CO2eq, with emissions per patient ranging from 25 to 2,452 kg CO2eq. Methodologically, the three most recent studies included nearly all emissions domains with high levels of data completeness, whereas the other three studies considered fewer than half of the emission domains, with medium to low data completeness. Only two studies fully disclosed their conversion factors. Four expert groups agreed on the need to develop standardized estimation tools for prospective use. Experts unanimously called for the involvement of all research stakeholders in raising global awareness of the carbon footprint of clinical trials.

**Interpretation:** The carbon footprint of clinical trials shows substantial variability, primarily due to differences in methodology and the domains of emissions assessed. Addressing these methodological inconsistencies with standardized and openly accessible tools is essential for developing strategies to reduce the environmental impact of clinical research, aligning with broader global efforts to mitigate climate change.

**Funding:** No funding

**Panel: Research in context:** *Evidence before this study:* Prior to this review, evidence on the carbon footprint of clinical research was sparse and inconsistent. A few opinion papers had briefly summarized the existing literature, but no formal review had been conducted. With a growing number of analytical publications in recent years – utilizing diverse assessment methods and reporting varying emissions - it became necessary to conduct a systematic review to compare and evaluated these methods and findings. Additionally, key recommendations made by experts, which seemed to align on several points, needed to be formally summarized.

*Added value of this study:* This is the first systematic review to critically evaluate and compare methodologies for estimating the carbon footprint of clinical trials. Our findings reveal wide variability in reported emissions, influenced by differences in study design, the emission domains assessed, the type of conversion factors used, and the reporting scale (by trial, by patient, or by year of execution). We emphasize the need for standardized, validated tools for consistent prospective carbon footprint assessments and advocate for the engagement of the research community to raise global awareness about this topic. This study lays the groundwork for advancing sustainable clinical research practices.

*Implications of all the available evidence:* The review highlights the importance of developing and adopting standardized tools for estimating the carbon footprint in clinical trials. These tools should be comprehensive, covering all relevant emission domains, and applied prospectively to support effective mitigation strategies from the start of the trial.

## Introduction

The climate footprint of the healthcare sector is substantial, contributing considerably to global greenhouse gas (GHG) emissions. In 2014, Pichler et al. estimated that the healthcare sector accounted on average for 5.5% of total national carbon emissions^1^. Specifically, it is estimated that the sector accounts for 8.5% of total emissions in the United States (US)^2^, 8.1% in France^3^, 6.3% in the United Kingdom (UK)^4^, 7% in Australia^5^, and 5% in Canada in 2018^6^. Major sources of GHG emissions include energy use in healthcare facilities, transportation, and the production and disposal of medical supplies^7^.

While the healthcare sector is essential for patient care, clinical research plays a crucial role in advancing healthcare practices. According to the definition proposed by the US National Cancer Institute (NCI)^8^, clinical research contributes to “finding new and better ways to detect, diagnose, treat, and prevent disease”, particularly through clinical trials. These trials are complex endeavors involving multiple stages with numerous activities in addition to standard care: ethical and regulatory protocol procedures, opening of investigating centers for patient inclusion, study execution with the research-specific care procedures, collection, storage, and transfer of data and samples (*e.g*., blood, tissues), monitoring, closing of centers, analysis and dissemination of results. Each stage involves various processes that may contribute to the overall carbon footprint of clinical trials.

Despite some institutional progress on climate issues since the early 2000s, including the UK’s Climate Change Act in 2008 and the Paris Agreement of 2015, research on the carbon footprint of clinical trials remains limited and inconsistent. In 2021, Adshead et al. estimated that the 350,000 trials registered on clinical ClinicalTrials.gov would approximately emit 27.5 million tons of CO2e^9^. This is equivalent to the annual CO2 emission of the city of Paris^10^. Given that these are global estimates, smaller-scale assessments are necessary to accurately quantify the carbon footprint of individual clinical trials and to design effective mitigation strategies by identifying the most carbon-intensive activities. A comprehensive review of existing methodologies and published results is equally important to avoid duplication, facilitate the dissemination of standardized and validated tools where available, or establish a foundation for developing new ones.

This study aims to synthesize the current evidence on the assessment of the carbon footprint of clinical trials. We specifically reviewed the methodologies used for carbon footprint estimation, including the scope of studies, data collection methods, and analytical processes.

## Methods

### 1) Search strategy and selection criteria

The present systematic review was conducted following the PRISMA 2020 guidelines^11^. The protocol was registered on PROSPERO (CRD42024569858, 08/05/2024). We searched Pubmed, Embase, and Cochrane databases with a research equation combining terms regarding clinical research (“clinical trials” OR “clinical research” OR “trial*”) and environmental footprint (“carbon footprint” OR “carbon emission*” OR “climate footprint” OR “environmental sustainability” OR “life-cycle analysis” OR “life cycle analysis” OR “environmental impact” OR “greenhouse effect” OR “greenhouse gas emission”). The research question was independently peer-reviewed using the PRESS 2015 checklist^12^ (provided in Appendix 1). Articles of any type, written in English and published from inception until April 16th, 2024, the date on which the selection was performed, were eligible for inclusion. For abstracts, further research was based on the authors’ names, title and publication to retrieve the full text. Abstracts without full-text availability were excluded from the analyses.

Two authors (CL, RL) independently screened the titles and abstracts to identify articles that required full-text examination, with a discussion on discordant cases until a consensus was found. Then, four authors in pairs (CL/CJ and ML/RL) independently examined full texts, extracted data, and discussed discordant cases until a consensus occurred. The references of included articles were checked to include potentially missed studies.

### 2) Data extraction

For each included study, the following data were extracted independently by four authors in pairs (CL/CJ and ML/RL): type of the study, year of publication, type of the trials evaluated (therapeutic area, type of intervention, phase and type of sponsoring), countries involved in the trials, duration of the trials, number of patients included in the trials. Details on the methodology implemented to evaluate the trials’ carbon footprint were also extracted: whether the tool used was described, validated in previous studies, included nine domains of emissions pre-identified using the referential published by the Low Carbon Clinical Trials Group from the Sustainable Healthcare Coalition^7,13^, and reasons for exclusion, when applicable. The emission domains encompassed all the activities of a clinical trial: i) trial set up (e.g., production of the trial documents and sending to trial sites, site activation,), ii) clinical trial unit (CTU) emissions (e.g., energy, heat, commuting to work), iii) trial-specific meetings and travel (e.g., travel to trial’s sites, accommodation), iv) treatment intervention (e.g. emissions associated with manufacturing, delivery, usage, and disposal of the trial’s therapeutic product), v) trial-specific patient assessment (emissions associated with medical procedures above the standard of care), vi) trial supplies and equipment (equipment used by investigators and patients specifically for the trial), vii) data collection and exchange (emissions associated to the collection, storage, and analysis of the trial data), viii) samples and lab (emissions associated with the collection, analysis, and storage of biological samples) and ix) trial close out (storage of trial documents, destruction of samples). Finally, reported results on carbon emissions for several functional units were extracted: total emissions by trial evaluated, average emissions by patient included, and by year of trial execution. The main carbon emitting domains were also retrieved for each trial evaluated.

### 3) Quality and biases estimation

The methodological quality of included studies was evaluated by the four authors during full-text screening following the Transparency Checklist for Carbon Footprint Calculations proposed by Lange et al^14^. This evaluation included the following items: specifications on the carbon emissions assessment boundaries, identification and description of the full life-cycle stages, description of the data sources (primary and/or secondary data) and extraction process, quality assessment of the conversion factors, temporal, geographical and technological representativeness of the conversion factors, reporting quality assessment and conduction of sensitivity analyses.

### 4) Data analyses

Articles included were classified into two categories: *analytical studies*, which aimed at quantifying the carbon footprint of one or several clinical trials; and *expert opinions* providing recommendations to structure and develop the global carbon emission assessment of clinical research.

#### Analytical studies

The characteristics of the evaluated trials, along with the carbon emission domains included in the evaluation and the resulting emissions were described using numbers (percentages) for categorical variables. Total emissions by trial were reported, and divided by the number of patients included to calculate individual emissions. We had initially planned to run a meta-analysis on the results and statistically compare the carbon emissions between similar trials. However, due to the limited number of studies included in this review and the heterogeneity of the trials evaluated, this meta-analysis could not be reasonably performed.

#### Expert opinion studies

The key messages and recommendations of these articles were analyzed, identified the recurrent themes and classified them into 5 recurrent categories.

The overall article selection process was performed using Zotero reference manager (version 6.0.36). Data extraction and analysis were conducted using Microsoft Excel.

## Results

A total of 1,048 studies were retrieved from PubMed, Embase, and Cochrane. After removing 335 duplicates, 713 publications remained for screening. Of these, 677 were filtered out based on titles, and 4 based on their abstracts. Additionally, 2 studies could not be retrieved and were therefore excluded. Thus, 30 articles remained for full-text review. Of these, 3 were only abstract papers and 15 were out-of-scope. This resulted in 12 studies being included in the final analysis: 6 analytical studies^7,15-19^ (Table 1) and 6 expert opinions^9,20-24^ (Table 2). The flowchart illustrating this selection process is displayed in Figure 1.

**Table 1.**
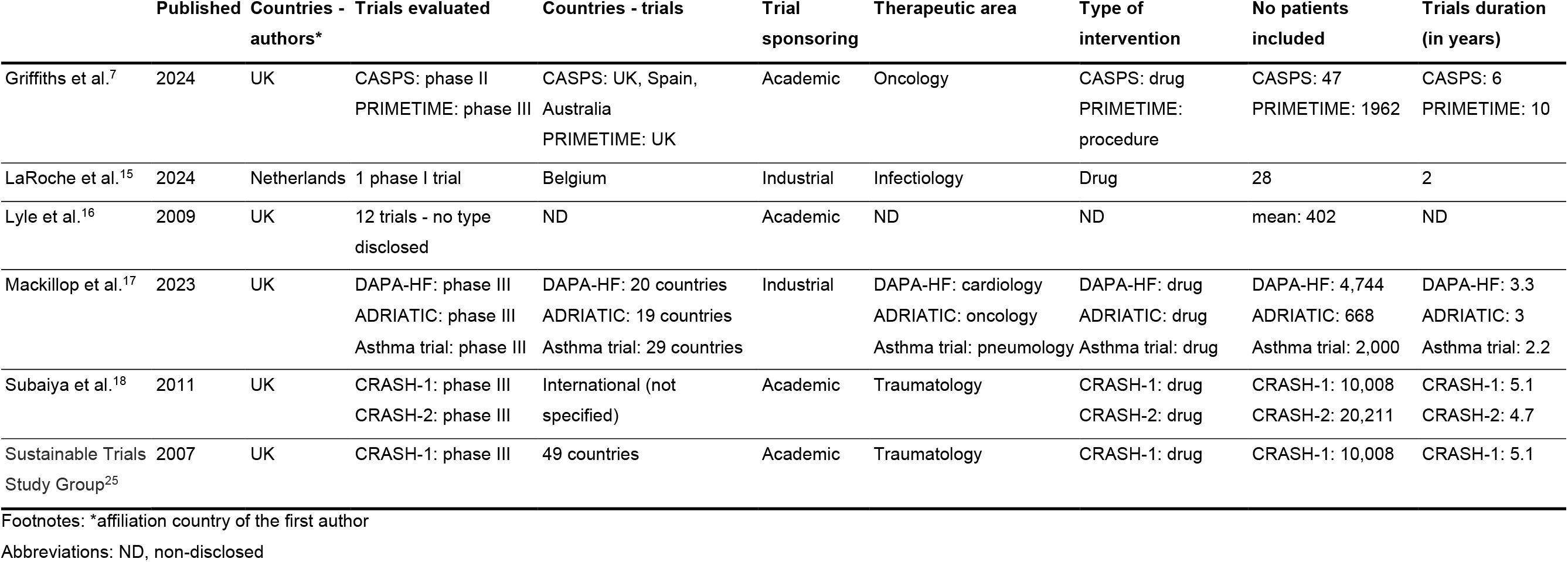
Description of the analytical studies included in this systematic review.

**Table 2.**
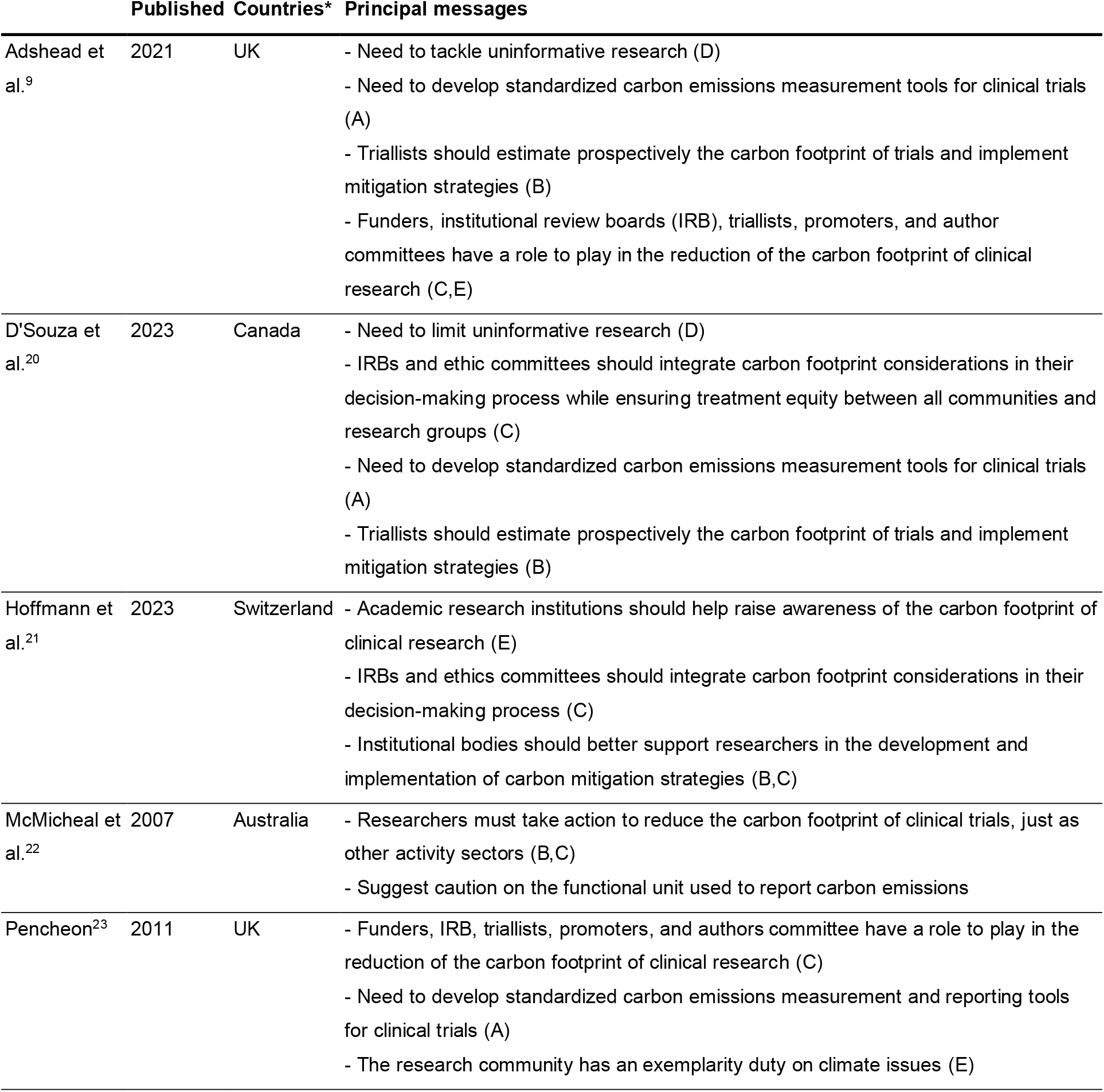
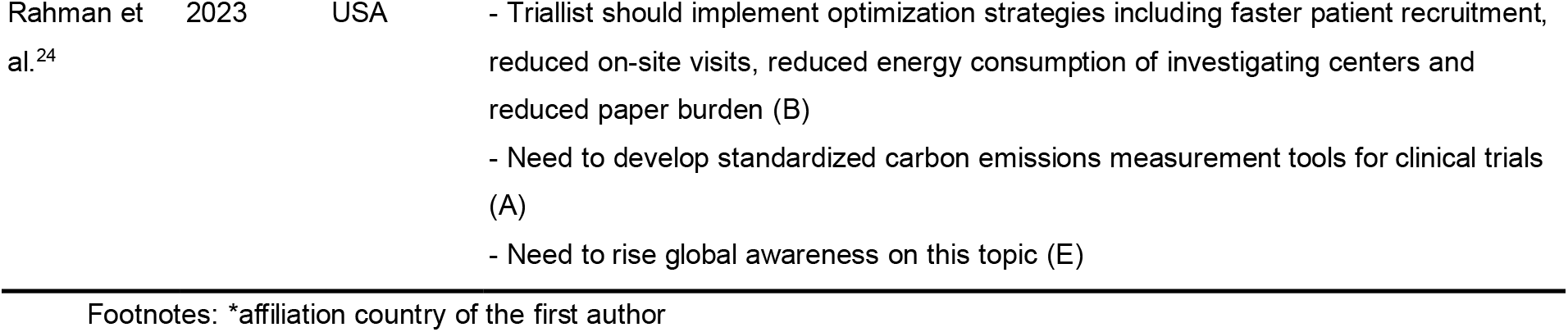
Description of the expert opinion articles included in this systematic review. Five recurring recommendations were identified from the expert opinion papers : A) The development of standardized estimation tools; B) The prospective estimation and mitigation of carbon emissions; C) The mobilization of all research stakeholders; D) The limitation of uninformative research and E) The need to raise global awareness on the carbon footprint of clinical trials.

**Figure 1.**
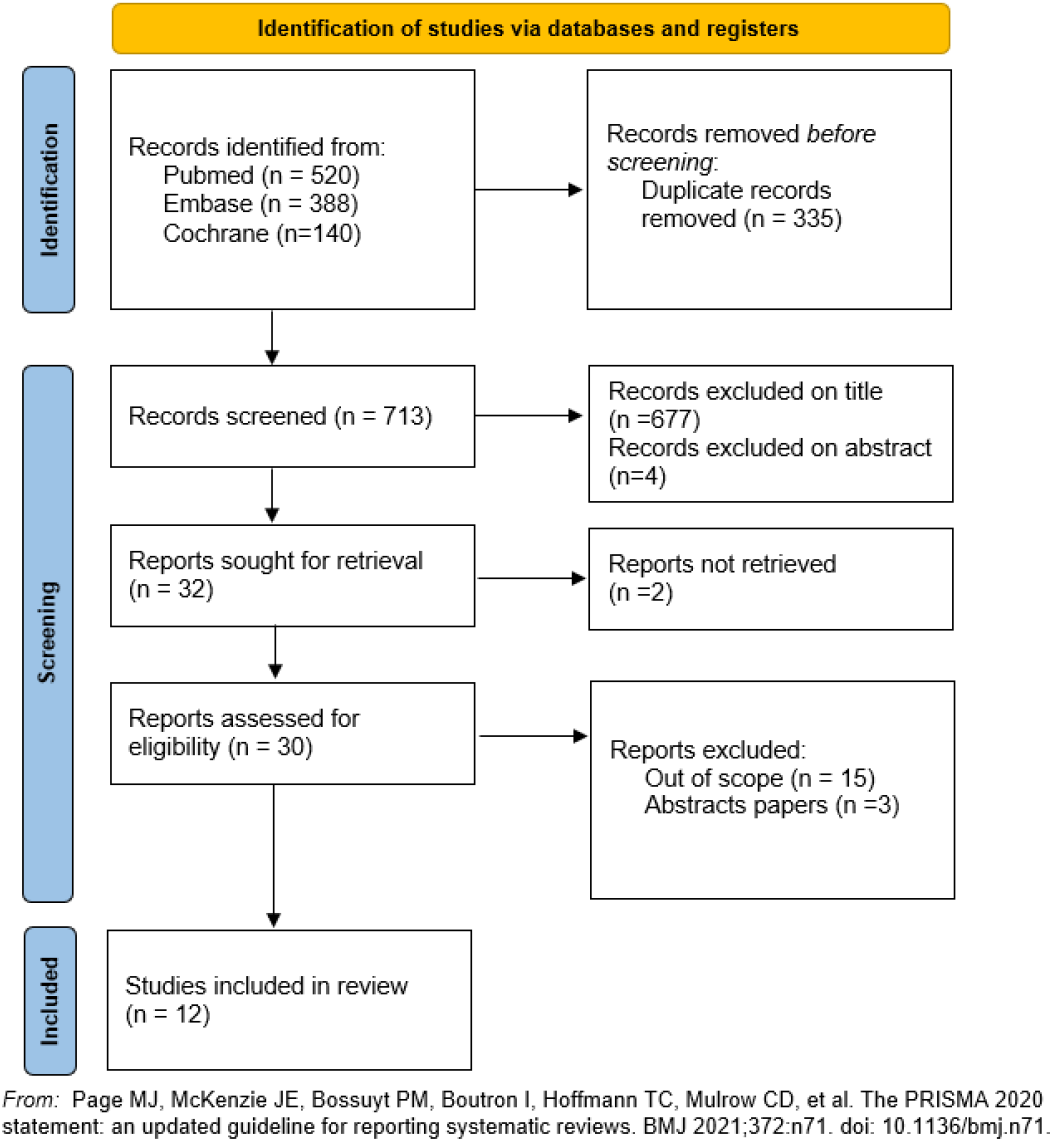
Study flowchart following the PRISMA 2020 guidelines.

The publication years of the included studies ranged from 2007 to 2024. Specifically, 3 analytical^16,18,25^ and 2 expert opinions^22,23^ were published up to 2011, and 7 additional studies (3 analytical^7,15,17^ and 4 expert opinions^9,20,21,24^) were published between 2021 and 2024. Out of the 12 studies, 7 (58.3%) were conducted in the UK^7,9,16-18,23,25^. The remaining studies were distributed across several other countries: one each in Australia^22^, Canada^20^, Netherlands^15^, USA^24^, and Switzerland^21^.

The analytical studies included a total of 20 clinical trials: one study included one industry-sponsored phase I trial^15^, two studies each included 2 academic trials (one phase I/one phase III and two phase III respectively)^7,18^, one study included 3 industry-sponsored phase III trials^17^, and one study incorporated 12 academic clinical trials (without specifying any trial information)^16^. Two studies^18,25^ evaluated the carbon footprint of the CRASH-1 trial. The clinical trials encompassed a diverse range of specialties, including oncology^17^, pneumology (asthma)^17^, cardiology (heart failure)^17^, infectiology (HIV/AIDS)^15^, traumatology^18,25^. All studied trials involved a pharmacological intervention, except for the PRIMETIME trial^7^ which evaluated a non-pharmacological strategy. The number of patients included in these trials markedly varied, running from 28 for the trial evaluated by LaRoche et al.^15^, up to 20,211 for the CRASH-2 trial^18^. The duration of the studies ranged from 2^15^ to 10^7^ years.

Total carbon emissions by trial and emissions by the included patients were displayed in Figure 2. Total emissions per trial ranged from 17.6 tons CO2eq for the trial evaluated by LaRoche et al.^15^ to 2,498 tons CO2eq for the DAPA-HF trial^17^. Emissions per patient also varied from 25 kg CO2eq for the CRASH-2 trial^18^ to 2,452 kg CO2eq for the ADRIATIC trial^17^. None of the studies reported other GHG emissions beyond CO2 equivalents. Notably, the two studies evaluating the carbon footprint of the CRASH-1 trial differed in reported emissions: Subaiya et al reported a total of 924.6 tons of CO2eq^18^ vs 642.6 tons of CO2eq for the study of the Sustainable Trials Study Group^25^.

**Figure 2.**
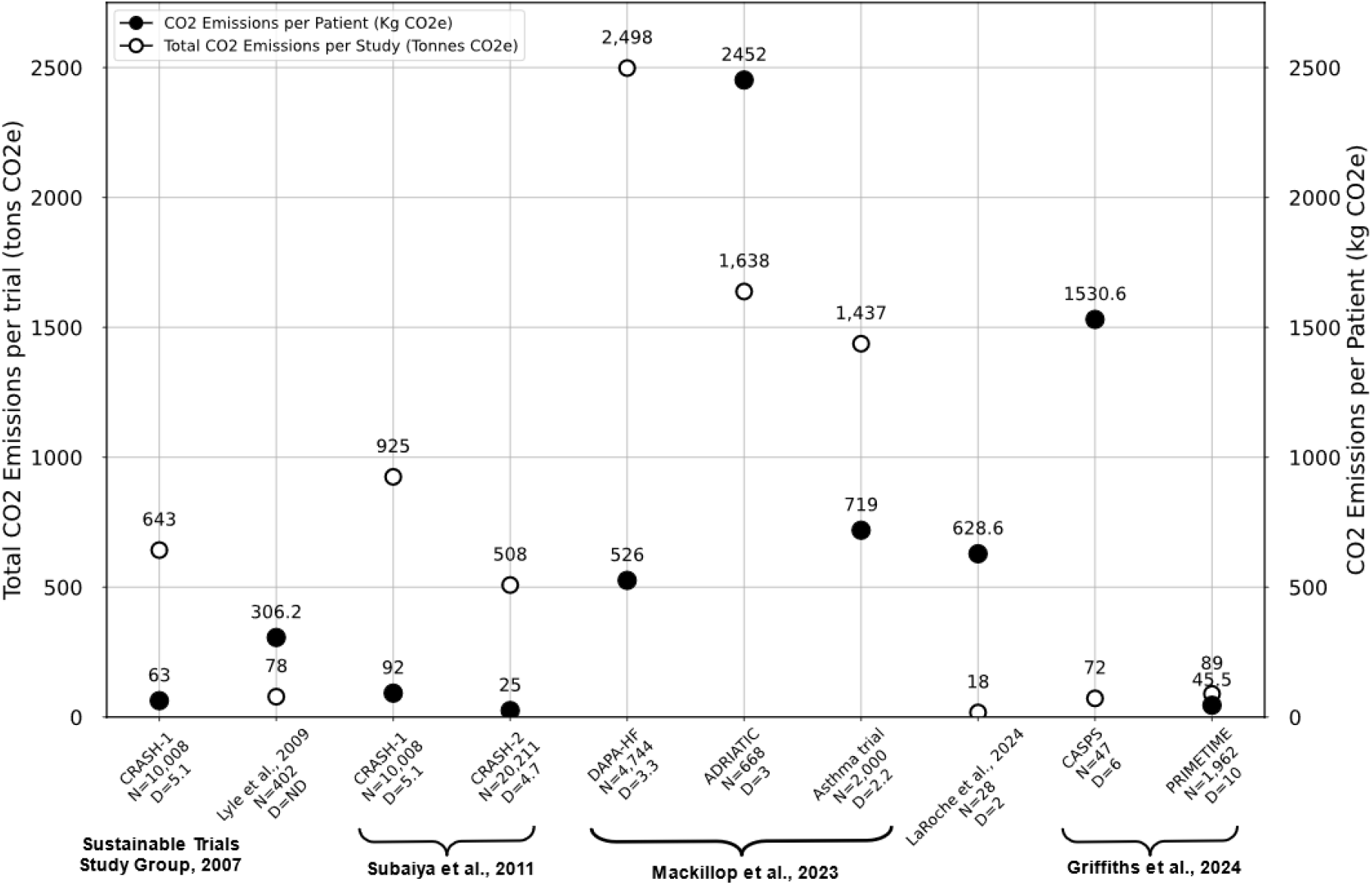
Total CO2e emissions per trial and emissions per patient. This figure compares total CO2 emissions per trial evaluated and corresponding emissions per patient included. The primary Y-axis (left) shows CO2 emissions in tons (Tons CO2e) per trial, while the secondary Y-axis (right) displays total CO2 emissions in kilograms (Kg CO2e) per patient. The X-axis lists the trials evaluated and their corresponding studies, including total patient count (N) and duration of the trial in years (D). Filled color-coded dots represent CO2 emissions per patient, and non-filled dots indicate total CO2 emissions per trial.

Methodologically, the three most recent studies tend to encompass a greater number of emission domains within their perimeter of analysis. Mackillop et al.^17^ and Griffiths et al.^7^ included all 9 carbon emission domains in their analysis (figure 3), while LaRoche et al.^15^ included 8 out of 9 domains (excluding the trial close out). In contrast, Lyle et al.^16^ and the Sustainable Trials Study Group^25^ included 4 domains (CTU emissions, trials-specific meetings, treatment intervention, trials supplies and equipment) and Subaiya et al.^18^ considered only CTU emissions, trial-specific emissions, and trial supplies and equipment. These discrepancies hinder the reporting and comparison of the highest-emitting domains across studies.

**Figure 3.**
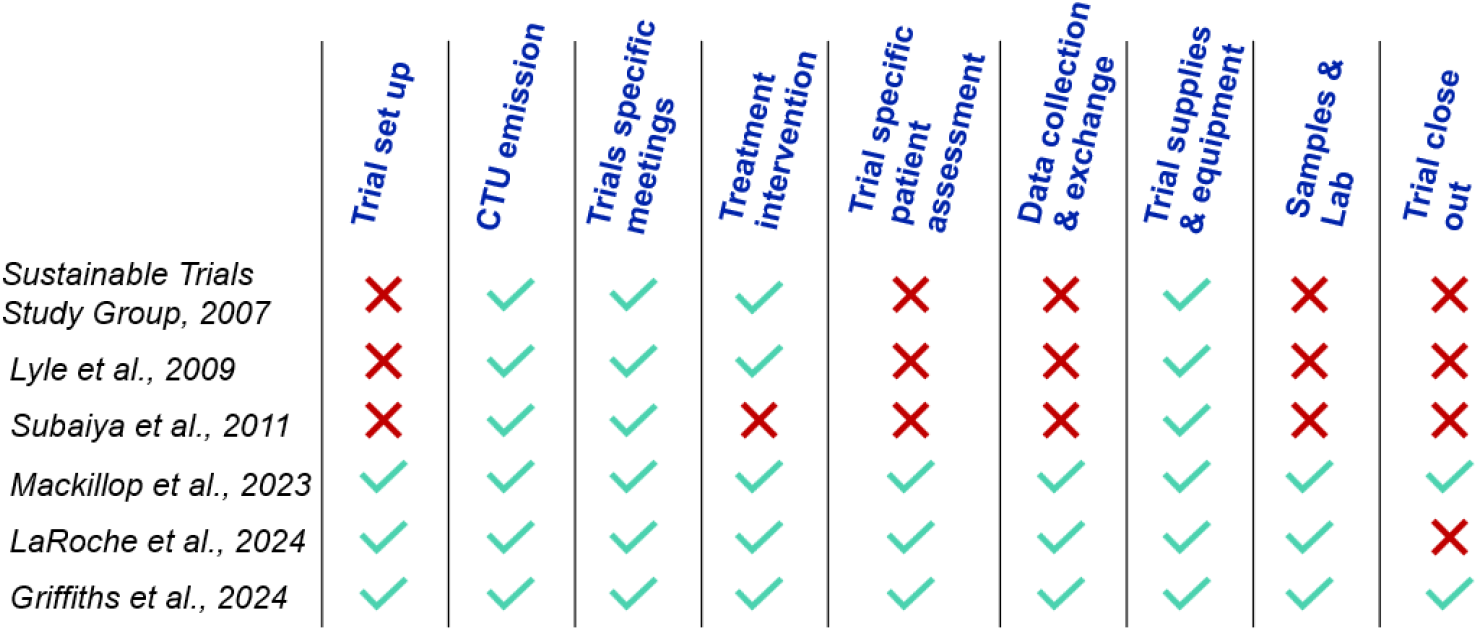
Carbon emission domains included in each analytical study. Emissions domains were retrieved from the life-cycle analysis proposed by Griffiths et al.^7^ and encompass all activities of a clinical trial.

We found variability in the quality of data used to evaluate the carbon footprint of clinical trials (Appendix 2). Specifically, two studies^7,16^ fully disclosed the conversion factors used to estimate emissions, while three studies^15,17,18^ partially published them (relying on both public and proprietary factors), and one study^25^ did not disclose any conversion factor. Furthermore, all studies utilized conversion factors that were found appropriate for the temporal, geographical, and technological context of their evaluations – except for the study of the Sustainable Trials Study Group^25^ which did not disclose their factors and for which evaluation was not possible. Finally, the reviewers estimated that data completeness of the carbon emission domains included was low in one study^25^, middle^16,18^ in two studies, and strong in three most recent studies^7,15,17^.

The systematic review identified six expert opinions on the carbon footprint of clinical trials. Experts converged on several key messages:

A. **Development of consensual estimation tools:** four articles^9,20,23,24^ advocated for creating standardized tools to estimate the carbon footprint of clinical trials. These tools should ensure consistency and comparability in carbon footprint measurements across different trials, providing a uniform basis for evaluating and addressing environmental impact.
B. **Prospective estimation and mitigation:** the importance of estimating the carbon footprint prospectively was highlighted in five articles^9,20-23^. This forward-looking approach would enable the implementation of mitigation strategies before trials begin, allowing researchers to address potential environmental impacts proactively.
C. **Mobilization of research stakeholders:** Experts uniformly highlighted the necessity of engaging a broad array of research stakeholders - funders, sponsors, ethics committees, trialists, and publishers. Their collective involvement is crucial for establishing a unified strategy to manage and reduce the carbon footprint of clinical trials.
D. **Limit uninformative research:** two articles^9,20^ insisted on avoiding duplicated research and urged triallists to double-check for previous publications, especially systemic reviews and meta-analyses, before starting any new research project.
E. **Raise global awareness:** four expert groups^9,21,23,24^ agreed on the importance of increasing global awareness about the carbon footprint of clinical trials, highlighting their critical contribution to the healthcare sector’s emissions. Raising awareness is essential to encourage further research on this topic, equip research stakeholders with the necessary background to critically assess future carbon emission estimates, and accelerate the implementation of mitigation strategies.

## Discussion

In this systematic review of the carbon emissions associated with clinical trials, we identified six analytical studies that assessed the carbon footprint of clinical trials and six expert opinions. Most studies (58.3%) were conducted by research teams in the UK. A total of 20 trials were evaluated, including one phase I trial, one phase II trial, six phase III trials and twelve trials with no reported information. The therapeutic areas of the evaluated trials included oncology, infectiology, cardiology, pneumology and traumatology. Except for one, all trials evaluated involved a pharmacological strategy. Reported carbon emissions varied widely from 17.6t CO2eq for the phase I trial evaluated by LaRoche et al.^15^ to 2,498t CO2eq for the DAPA-HF trial evaluated by Mackillop et al.^17^ Similarly, the methods used to estimate carbon emissions were inconsistent, with diverse domain emissions included and data completeness levels. Furthermore, four of the six included opinion papers emphasized the need for the development of standardized, internationally validated tools for measuring carbon emissions. Five expert groups called for the prospective use of these tools by investigators to limit the carbon footprint of trials from their inception. Experts unanimously praised for the broad mobilization of all clinical research stakeholders. Two articles stressed the importance of limiting uninformative research, and four emphasized the need to raise global awareness of the carbon footprint of clinical trials.

Several factors can influence the reporting of carbon emissions from clinical trials. Firstly, the number of emission domains considered, and the completeness of emissions measured within each domain greatly affect the overall reported emissions of a trial. We observed a trend in recent studies to include more emission domains and increase their data completeness^7,15,17^, leading to higher reported emissions by trial and by patient compared to older studies with a more limited carbon evaluation perimeter^16,18,25^. However, these studies provided a more accurate representation of the trials’ true emissions. Additionally, Subaiya et al.^18^ reported higher emissions for the CRASH-1 trial than the Sustainable Trials Study Group^25^, despite including fewer emission domains. This discrepancy can be attributed to differences in data completeness within each emission domain. For example, Subaiya et al.^18^ included 13t CO2e per year of natural gas emissions for CTU emissions, while the Sustainable Trials Study Group^25^ did not consider these. There were also differences in reported emissions from electricity consumption by the CTU (29t CO2e per year for Subaiya et al.^18^ vs 45.4t CO2e for the Sustainable Trials Study Group^25^), though the original articles did not provide sufficient methodological details to explain this difference. This underscores the need for standardized and publicly available measurement tools to facilitate comparison between studies. Secondly, the scale used to report emissions greatly influences the interpretation and comparison of trials. For instance, the DAPA-HF trial^17^ was the highest-emitting trial in terms of total global carbon emissions, yet it ranked fifth in terms of emissions by patient, whereas the trial evaluated by LaRoche et al.^15^ had the lowest overall emissions but ranked fourth highest in emissions by patient. This variability may be partly explained by differences in trial design: the trial studied by LaRoche et al.^15^ was a phase I trial involving only 28 patients, while the DAPA-HF^17^ phase III trial included 4,744 patients across 20 countries. The trials also differed in therapeutic area, with cardiology (DAPA-HF^17^) potentially requiring more medical devices and trial supplies than infectiology (LaRoche et al^15^). This suggests the need for caution when comparing the carbon emissions of clinical trials, taking into account the methodology used, the reporting scale employed by investigators, the type of trials and therapeutic areas being compared.

We observed discrepancies in the methodological approach of analytical studies depending on the type of sponsoring. Academic research teams evaluating trials often could not access data on the carbon emissions associated with drug manufacturing. As a result, they either estimated these emissions using financial conversion factors^16^ or excluded them from their study scope^7,18,25^, while still including emissions related to drug packaging, transport, and disposal. This is concerning, especially since emissions from drugs and medical devices are prominent contributors to the overall carbon footprint of healthcare systems^2-4^. In contrast, industry-sponsored researchers had access to manufacturing data, allowing them to quantify the associated emissions^15,17^. However, they had limited access to information on clinical activities involved in the evaluated trials and thus relied on secondary estimates from trial records. This highlights the pressing need for collaboration between drug manufacturers and academic research teams to facilitate access to comprehensive data for each process of the trials, ensuring the most accurate estimations possible. Furthermore, nearly all trials evaluated were pharmacological, and trials investigating non-pharmacological strategies and devices should be given the same consideration as pharmacological trials when estimating carbon footprints. Future research must ensure that all types of trials are evaluated.

As highlighted in several opinion papers^9,20,21,23^, the estimated carbon emissions of future clinical trials will need to be considered in funding applications and ethical approvals, along with mitigation strategies to limit their environmental footprint. While this will aid granting committees in deciding between comparable trial designs and therapeutic areas, it should remain a secondary criterion, behind the clinical impact of research. Large phase III studies, which inherently produce high global emissions due to their design and the necessity of including many patients from multiple centers, should not receive fewer grants because of their total emissions. Similarly, research on rare diseases, which demands extensive research and development phases for a small number of patients, should not be disadvantaged due to their high carbon emissions per patient. As with financial aspects, carbon emissions should be used to compare similar trial designs and therapeutic areas.

Given the urgency of the global climate crisis, and as emphasized by Pencheon^23^, the healthcare sector has an exemplarity duty and must embark on a new path of sustainable healthcare that combines sobriety, efficiency, and mitigation strategies. All stakeholders, including the clinical research community, must unite in this effort. Accurate quantification of the sector’s carbon footprint, at both small and large scales, is crucial for identifying the most carbon-intensive areas and guiding transformation strategies. If interest in evaluating the environmental impact of clinical research began 20 years ago, spurred in the UK by the Climate Change Act., only five studies were published between 2007 and 2011^16,18,22,23,25^, followed by a 10-year gap in the scientific literature. Recently, there has been a resurgence of interest, with seven studies published since 2021 by international research teams^7,9,15,17,20,21,24^. Notably, the Low Carbon Clinical Trials (LCCT) working group was established within the UK Sustainable Healthcare Coalition to develop strategies for reducing the carbon footprint of clinical trials. They have created and published an openly accessible measurement tool for academic trials (https://clinicaltrialcarbon.org/ - beta version), currently under validation in the UK. Sharing methodologies and conversion factors is essential to ensure the comparability and transparency of estimates, while also enabling their adaptation to the specificities of each country and domain. Most importantly, this broad mobilization requires awareness campaigns, from academic courses to lifelong training, to engage as many actors as possible and prioritize sustainable research and healthcare in their routine practice.

Our study has several strengths. Firstly, to the best of our knowledge, this is the first systematic review focusing on the evaluation of the carbon footprint of clinical trials. While several expert opinion papers have drawn up observations and made some recommendations, no formal assessment has been conducted until now. Secondly, our systematic and comprehensive review of the methodologies and tools used to estimate the carbon footprint of trials highlighted the caution required when comparing reported emissions between trials and underscored the importance of using standardized, internationally validated tools. This study will help document the evolution of research in this field and contribute to the research community’s mission to disseminate its findings^26^ and raise awareness about its carbon footprint. It also aims to be repeated in the future, allowing for a formal comparison of carbon emissions between trials.

However, our study also has limitations. First, statistical pooling and comparison of the carbon emissions between trials were not feasible due to the limited number of analytical studies and the low comparability between trials, which varied in phases, settings, and therapeutic areas. Second, we focused only on carbon emissions, not on the broader environmental footprint of clinical trials. While the carbon footprint is one of the most documented and quantified planetary boundaries, guiding our choice, the impact of clinical research on biodiversity, water pollution, and other greenhouse gases should be considered.

## Conclusion

This systematic review highlighted the urgent need for standardized method to assess the carbon footprint of clinical trials and identify areas for mitigation and improvement. The healthcare sector, while focused on improving health and care, significantly contributes to greenhouse gas emissions, creating a feedback loop that worsens climate change and public health. Integrating sustainability into clinical research is crucial to breaking this cycle and aligning healthcare with global climate efforts.

## Contributors

All the authors participated in the conceptualization of this study. CJ ran the online search, and CL and RL screened titles and abstracts. CJ, CL, RL, and ML performed the full-text review and data extraction. ML did the data visualization. CJ and ML proposed the original manuscript draft, and all the authors contributed to the review and editing process before submission.

## Data Availability

All data produced in the present work are contained in the manuscript

## Data sharing

No personal data was collected for this study.

## Acknowledgments

No acknowledgement.

