## Supplemental Materials for "Assessing the carbon footprint of clinical trials: a systematic review"

#### Supplementary materials

##### Appendix 1. PRESS checklist

***PRESS Guideline* — Search Submission & Peer Review Assessment**

**SEARCH SUBMISSION: THIS SECTION TO BE FILLED IN BY THE SEARCHER**

| | |
|---|---|
| Searcher: Charline JEAN | |
| Date submitted: 25/07/2024 | Date requested by: 26/07/2024 |

**Systematic Review Title:**

Evaluating the carbon footprint of clinical trials: a systematic review

This search strategy is ...

| | |
|---|---|
| X | My PRIMARY (core) database strategy — First time submitting a strategy for search question and database |
| | My PRIMARY (core) strategy — Follow-up review NOT the first time submitting a strategy for search question and database. If this is a response to peer review, itemize the changes made to the review suggestions |
| | SECONDARY search strategy— First time submitting a strategy for search question and database |
| | SECONDARY search strategy — NOT the first time submitting a strategy for search question and database. If this is a response to peer review, itemize the changes made to the review suggestions |

**Database**

(i.e., MEDLINE,CINAHL…): *[mandatory]*

PubMed, Embase and Cochrane

**Interface**

(i.e., Ovid, EBSCO…): *[mandatory]*

None

**Research Question**

(Describe the purpose of the search) *[mandatory]*

We aimed to synthetize current evidence on the evaluation of the carbon footprint of clinical trials. We specifically reviewed the methodologies of carbon footprint estimation, including the scope studied, data collection and analytic processes.

#### PICO Format

(Outline the PICOs for your question — i.e., Patient, Intervention, Comparison, Outcome, and Study Design — as applicable)

| | |
|---|---|
| P | Clinical trials of any type and any stage |
| I | Activities related to the clinical trials |
| C | Not applicable, as carbon emissions will be compared between included |
| O | Reported carbon emissions by clinical trial |
| S | Studies of any type |

#### Inclusion Criteria

(List criteria such as age groups, study designs, etc., to be included) *[optional]*

Articles in english, of any type, published from inception until 16/04/2024, were eligible for inclusion.

Articles were excluded from the analyses if they were out of scope. In case of abstracts, further research based on the authors' names, title and publication to retrieve full text, abstracts alone were excluded from the analyses

#### Exclusion Criteria

(List criteria such as study designs, date limits, etc., to be excluded) *[optional]*

None

#### Was a search filter applied?

Yes ☐ No ■

**If YES, which one(s) (e.g., Cochrane RCT filter, PubMed Clinical Queries filter)? Provide the source if this is a published filter.** *[mandatory if YES to previous question — textbox]*

Other notes or comments you feel would be useful for the peer reviewer? ***[optional]***

Please copy and paste your search strategy here, exactly as run, including the number of hits per line. ***[mandatory]***

**<u>For Pubmed (520 hits)</u>**: (

("clinical trials as topic"[MeSH Terms]) OR ("clinical trial*"[Text Word]) OR ("clinical research"[Text Word]) OR ("trial*"[Text Word])

) AND (

("carbon footprint"[MeSH Terms]) OR ("carbon footprint"[Text Word]) OR ("carbon emission*"[Text Word]) OR ("climate footprint"[Text Word]) OR ("environmental sustainability"[Text Word]) OR ("life cycle analysis"[Text Word]) OR ("life-cycle analysis":ti,ab,kw) OR ("environmental impact"[Text Word]) OR ("greenhouse effect"[Text Word]) OR ("greenhouse gas emission"[Text Word])

)

**<u>For Embase (388 hits)</u>** : (

("clinical trial*":ti,ab,kw) OR ("clinical research":ti,ab,kw) OR ("trial*":ti,ab,kw)

) AND (

("carbon footprint":ti,ab,kw) OR ("carbon emission*":ti,ab,kw) OR ("climate footprint":ti,ab,kw) OR ("environmental sustainabilitiy":ti,ab,kw) OR ("life cycle analysis":ti,ab,kw) OR ("life-cycle analysis":ti,ab,kw) OR ("environmental impact":ti,ab,kw) OR ("greenhouse effect":ti,ab,kw) OR ("greenhouse gas emission":ti,ab,kw)

)

**<u>For Cochrane (140 hits)</u>** : (

("clinical trial*"):ti,ab,kw OR ("clinical research"):ti,ab,kw OR (trial*):ti,ab,kw

) AND (

("carbon footprint"):ti,ab,kw OR ("carbon emission*"):ti,ab,kw OR ("climate footprint"):ti,ab,kw OR ("environmental sustainabilitiy"):ti,ab,kw OR ("life-cycle analysis"):ti,ab,kw OR ("life cycle analysis"):ti,ab,kw OR ("environmental impact"):ti,ab,kw OR ("greenhouse effect"):ti,ab,kw OR ("greenhouse gas emission"):ti,ab,kw

)

**(Add more space, as necessary.)**

#### PEER REVIEW ASSESSMENT: THIS SECTION TO BE FILLED IN BY THE REVIEWER

| Reviewer:Hédi Chabanol | Date completed:12/09/2024 |
|---|---|

##### 1. TRANSLATION

| | |
|---|---|
| A --- No revisions | X |
| B --- Revision(s) suggested | ☐ |
| C --- Revision(s) required | ☐ |

If "B" or "C," please provide an explanation or example:

##### 2. BOOLEAN AND PROXIMITY OPERATORS

| | |
|---|---|
| A --- No revisions | X |
| B --- Revision(s) suggested | ☐ |
| C --- Revision(s) required | ☐ |

If "B" or "C," please provide an explanation or example:

##### 3. SUBJECT HEADINGS

| | |
|---|---|
| A --- No revisions | X |
| B --- Revision(s) suggested | ☐ |
| C --- Revision(s) required | ☐ |

If "B" or "C," please provide an explanation or example:

##### 4. TEXT WORD SEARCHING

| | |
|---|---|
| A --- No revisions | X |
| B --- Revision(s)suggested | ☐ |
| C --- Revision(s) required | ☐ |

If "B" or "C," please provide an explanation or example:

##### 5. SPELLING, SYNTAX, AND LINE NUMBERS

| | |
|---|---|
| A --- No revisions | X |
| B --- Revision(s)suggested | ☐ |
| C --- Revision(s) required | ☐ |

If "B" or "C," please provide an explanation or example:

##### 6. LIMITS AND FILTERS

| | |
|---|---|
| A --- No revisions | X |
| B --- Revision(s) suggested | ☐ |
| C --- Revision(s) required | ☐ |

If "B" or "C," please provide an explanation or example:

OVERALL EVALUATION (Note: If one or more "revision required" is noted above, the response below must be "revisions required".)

| | |
|---|---|
| A --- No revisions | X |
| B --- Revision(s) suggested | ☐ |
| C --- Revision(s) required | ☐ |

Additional comments:

### Appendix 2. Risk of bias assessment

| | Assessment boundaries detailed | Life-cycle stages described | Type of data sources (primary and/or secondary) | Publication of the CF | CF temporal representativeness | CF spatial representativeness | CF technological representativeness | Data completeness |
|---|---|---|---|---|---|---|---|---|
| Griffiths et al (2024) | Yes | Yes | Mostly primary | Yes | Yes | Yes | Yes | High |
| LaRoche et al (2024) | Yes | Yes | Mixed | Partially | Yes | Yes | Yes | High |
| Lyle et al (2009) | Partially | Partially | Mostly secondary | Yes | Yes | Yes | Yes | Middle |
| Mackillop et al (2023) | Yes | Yes | Mixed | Partially | Yes | Yes | Yes | High |
| Subaiya et al (2011) | Partially | Partially | Mostly secondary | Partially | Yes | Yes | Yes | Middle |
| Sustainable Trials Study Group (2007) | No | No | Mostly secondary | No | NA | NA | NA | Low |

Footnotes: *affiliation country of the first author

Abbreviations: CF, conversion factors; NA, not-applicable

This risk of bias assessment was based on the Transparency Checklist for Carbon Footprint Calculations proposed by Lange et al.[14]

The temporal representativeness assesses whether the publication period of the conversion factors used is relevant to the trial evaluated (e.g., whether the conversion factors vary with temporal changes in the energy mix of countries). The spatial representativeness assesses whether the geographical location of the data used to emit conversion factors is relevant to the trial evaluated (e.g., whether the conversion factors associated with electricity production depend on the country of the trial evaluated). The technological representativeness assesses whether the technology used to emit conversion factors is relevant to the trial evaluated (e.g., for transport, whether different conversion factors are used depending on the type of transport – plane, train, bike).

Data completeness was subjectively assessed by reviewers in pairs (CJ/CL and RL/ML) depending on the overall level of completeness within emission domains. Discordant cases were discussed until a consensus was reached.
